# Remdesivir for COVID-19: match-population analysis with compassionate use of Remdesivir for severe COVID-19

**DOI:** 10.1101/2020.07.22.20160002

**Authors:** Olga Vasylyeva, Tara Chen, John Hanna

## Abstract

We retrospectively compared outcomes between patients who received compassionate care Remdesivir and whose who met criteria for Remdesivir, but did not received it due to period of unavailability. We observed comparable mortality rate and significantly higher mortality rate among patients with CrCl < 30 ml per minute.

## Background

COVID-19 is a disease caused by SARS-CoV2 coronavirus that can result in severe acute respiratory syndrome and is classified as a pandemic by the World Health Organization (WHO). [1,2] As of July, 2020, in the United States, 140,828 people died due to COVID-19 that is the highest mortality among other countries. [3]

Up to date, no U.S. Food and Drug Administration (FDA) approved antiviral COVID-19 drugs that demonstrated efficacy in a completed randomized controlled trial is available. On May 1, 2020, the FDA issued emergency use authorization for investigational drug Remdesivir in hospitalized patients with severe COVID-19. [4-7]

Remdesivir is an experimental nucleoside analogue developed by Gilead Sciences that works by blocking viral RNA synthesis. It was reported that Remdesivir has activity against MERS, SARS, and SARS-CoV2 *in vitro*. [8] Since January 2020, Gilead Sciences accepted requests for compassionate use Remdesivir, however on March 22, 2020 Gilead Sciences introduced limitation on compassionate use except for pregnant women and children due to overwhelming demand. [9, 10]

On April 10, 2020, multicenter data were published by Grein J. et al. on compassionate use of Remdesivir for patients with severe COVID-19 based on the FDA and Gilead Sciences approval before the restriction, but the study lacked a control group. [10] Rochester Regional Health (RRH) has a number of hospitalized COVID-19 patients who did not receive Remdesivir due to its unavailability. We attempted to match the population of patients hospitalized to RRH for COVID-19 after the introduction of restriction with the patients’ population who received Remdesivir for compassionate use as reported by Grein J. et. al.

## Method

This is a retrospective review of the patients’ data who was hospitalized due to COVID-19 since March 22, 2020. Patients were selected to match inclusion criteria to receive compassionate care Remdesivir as reported by Grein J. et al. and Gilead Science. Inclusion criteria were: hospitalized patients with laboratory-confirmed SARS-CoV2 by polymerase chain reaction (PCR); oxygen saturation of 94% or less while the patient was breathing ambient air or a need for oxygen support; creatinine clearance (CrCl) > 30 ml per minute upon admission; aspartate aminotransferase (AST) and alanine aminotransferase (ALT) less than five times the upper limit of normal range upon admission. The primary outcomes were a need for invasive ventilation and mortality.

## Results

Between 3/22/2020 and 4/30/2020, 149 patients with laboratory confirmed COVID-19 were hospitalized to RRH. Among these patients, 98 (65.7 %) met inclusion criteria for compassionate use of Remdesivir either at the time of admission or during hospitalization.

Median age was 64 (27-97) years, and 62 (63.2%) were males. Median time of admission was eight days. Forty (40.8 %) patients required intensive care unit admission, and 20 (20.4 %) invasive ventilation. Ten patients died (10.2 %) with median time from admission to death nine days. (Table 1)

**Table 1.** Baseline characteristics and outcomes of patients received and did not receive compassionate care Remdesivir.

| **Baseline characteristics and Outcomes** | **Received compassionate care Remdesivir (Grein J. et al.)** | **Remdesivir not available (RRH)** |
| --- | --- | --- |
| Total number of patients meeting criteria for compassionate use of Remdesivir | 53 | 98 |
| Median age, yr | 64 | 64 |
| Male sex, n (%) | 40 (75%) | 62 (63.2 %) |
| Duration of hospitalization, median | Unknown | 8 days |
| Invasive ventilation, n (%) | 34 (64%) | 20 (20.4%) |
| Death, n (%) | 7 (13%) | 10 (10.2%) |

Among 51 patients who was not qualified to receive Remdesivir based on inclusion criteria, 22 had CrCl < 30 ml per min upon admission. Eight out of these patients (36.3 %) required invasive ventilation, and eight patients died, which was significantly more than patients with CrCl > 30 ml min (36.3% vs 10.2%, *p* = 0.001). Five out of 51 were not eligible for Remdesivir due to elevated AST and ALT more than five times of normal limits upon admission.

## Limitations

The major limitation of this study is the lack of randomization and sufficient matching to perform statistical analysis on difference as we do not have raw data from the group that received Remdesivir. Out population had fewer males (63.2% vs. 75%) and notably fewer patients requiring mechanical ventilation (20.4% vs. 64%). The median age in both groups was the same, 64 years.

Six patients in the group that did not receive Remdesivir remained hospitalized at the time of follow up period, and final outcome is unknown.

## Discussion

Grein J. et al. article on outcomes for the patients who received Remdesivir for compassionate care reported 13% mortality, but there was no control group. We compared this data with the population who met the same criteria for compassionate care Remdersivir, but did not receive it because of unavailability.

In our population, 10 (10.2%) patients died. It is unknown if there is a statistical difference between groups as we have no detailed data from the group who received Remdesivir. We observed that in our population, much fewer people required invasive ventilation (20.4% vs. 64%). Furthermore, patients who presented with CrCl < 30 per min had significantly higher mortality as compared with patients who met criteria to receive Remdesivir (36.3% vs. 10.2%). Benefits of Remdesivir for patients required invasive ventilation and have CrCl < 30 per min require further investigation.

On May 22 2020, a preliminary report from a clinical trial comparing Remdesivir and placebo for treatment of COVID-19 was published [11,12]. Preliminary results showed 10.7% mortality in the placebo group, which is comparable with 10.2% in our population, and 5.9% in Remdesivir group. In this clinical trial, the difference in mortality between placebo and intervention groups was not statistically significant. Median time to recovery, defined in the trial as either discharge or continuous hospitalization for infection-control purposes, significantly decreased from 15 to 11 days. However, in our population median time of hospitalization was eight days, therefore it is uncertain if we will be observing this benefit in our population. The final report of clinical trial is presently pending.

In conclusion, mortality among people who met inclusion criteria for compassionate use Remdesivir but did to received it due to unavailability was 10.2%. There was significantly higher mortality among patients presented with CrCl < 30 ml per min, an exclusion criterion for compassionate use of Remdersivir, and the benefit of Remdesivir among this population needs to be further evaluated.

## Data Availability

There is no external dataset or supplementary material online are available. Institutional approval is required for any protected data as per IRB.

## Funding

This work was not supported by any funding.

## Conflict of Interest

None.

## Notes

### Competing Interest Statement

The authors have declared no competing interest.

### Funding Statement

No funding.

### Author Declarations

Office of Human Research Protection at Rochester Regional Health. IRB 1992.

